# Body composition and melanoma incidence risk: insights from a longitudinal lung cancer screening cohort

**DOI:** 10.1101/2025.10.10.25337772

**Authors:** Tong Yu, Grant Kokenberger, Jing Wang, Xin Meng, Diwakar Davar, Walter J. Storkus, John M. Kirkwood, Hassane M. Zarour, Jiantao Pu

**Affiliations:** Department of Radiology, University of Pittsburgh, Pittsburgh, PA 15213, USA; Department of Bioengineering, University of Pittsburgh, Pittsburgh, PA 15213, USA; Division of Hematology-Oncology, Department of Medicine, University of Pittsburgh, Pittsburgh, PA 15213, USA; Department of Dermatology, University of Pittsburgh, Pittsburgh, PA 15213, USA

**Keywords:** melanoma, body composition, competing risk, opportunistic screening

## Abstract

**Objective:** This study explored the association between low-dose computed tomography (LDCT)-derived body composition and melanoma incidence risk.

**Methods:** LDCT scans from the Pittsburgh Lung Screening Study (n=3,422, 22 follow-up years) were analyzed. Body composition features were segmented and quantified from baseline scans using in-house artificial intelligence algorithms. Features were selected before modeling. Fine-Gray subdistribution hazard models assessed the association between body composition and melanoma incidence. Model performance was evaluated using time-dependent area under the curve (AUC). Restricted mean survival time (RMST) compared melanoma-free survival across BMI and body composition groups at 5, 10, and 15 years. Participants were stratified into risk groups, with risk estimated at each time point. Sex-specific analyses were conducted separately. Statistical significance was defined as p<0.05.

**Results:** Among 3,422 participants, 80 developed melanoma (43 males, 37 females). In the overall model, visceral adipose tissue (*VAT*) *volume* (hazard ratio [HR]=1.27), skeletal muscle (*SM*) *density* (HR=0.81), and *bone density* (HR=1.33) were included, achieving a 21-year AUC of 0.68 (95% CI: 0.65–0.70). The male-specific model included only *SM density* (HR=0.74; AUC=0.67, 95% CI: 0.65–0.68). The female-specific model (AUC=0.68, 95% CI: 0.65–0.71) included *VAT volume* (HR=1.47), intramuscular adipose tissue (*IMAT*) *ratio* (HR=0.67), and *bone density* (HR=1.75). Higher *VAT, IMAT volume*, and lower *SM density* showed shorter melanoma-free survival and stratified risk better than BMI. Males exhibited higher estimated risk than females.

**Conclusion:** LDCT-derived body composition metrics may provide incidental insights into melanoma risk during lung cancer screening, though their predictive utility remains limited and warrants further investigation.

**Key Points:** *Question:* To investigate the association between CT-derived three-dimensional (3D) body composition and the risk of developing melanoma.

*Findings:* CT-derived body composition was associated with melanoma incidence. Males demonstrated higher estimated risk than females over both short- and long-term follow-up periods.

*Clinical Relevance:* Given melanoma’s high mortality and the limited effectiveness of current screening programs, these findings highlight the potential of leveraging routinely acquired lung cancer screening CT scans to enhance melanoma risk assessment.

## 1. Introduction

Melanoma is the fifth most common cancer among both men and women in the United States [1], responsible for approximately 75% of skin cancer-related deaths despite representing only 4% of cases [2]. In 2024, an estimated 100,640 new cases and 8,290 deaths were expected in the U.S. [1]. Early detection relies on self-examinations, clinical skin exams, and dermatologic evaluations, which focus on identifying cutaneous abnormalities or changes in mole size, shape, or color. Confirmatory diagnosis is established through histopathological analysis of suspicious lesions [3]. While various screening programs have been proposed in the U.S. to identify melanoma in at-risk adults to reduce mortality [4], the U.S. Preventive Services Task Force found insufficient evidence to recommend routine clinical skin screenings for early detection [5]. Consequently, no standard screening protocol exists for the general population [6]. Additionally, screenings often detect benign lesions, such as nevi, that may never progress to invasive melanoma, while some early-stage melanomas may be missed [7]. These limitations contribute to false positives and negatives, resulting in unnecessary follow-ups or missed diagnoses, emphasizing the need for alternative approaches to complement existing methods and improve early detection.

Low-dose CT (LDCT) scans, widely adopted for early lung cancer screening in the U.S., provide high-resolution imaging that enables detailed evaluation of various anatomical structures. Beyond their primary purpose, LDCT scans have spurred interest in opportunistic screening (OS) [8, 9]. By repurposing routine clinical imaging data, OS offers a cost-effective and resource-efficient alternative to dedicated screening programs, maximizing the utility of already-acquired images [10]. Enhanced with artificial intelligence (AI), LDCT-based OS has shown promise in detecting unsuspected presymptomatic diseases [9], such as osteoporosis [11], sarcopenia, cardiovascular disease, and diffuse liver diseases [8], primarily through quantitative body composition analysis.

Body composition characteristics, such as visceral adiposity and sarcopenia, are increasingly recognized as modulators of cancer biology. In melanoma, these characteristics may influence tumor initiation and progression through chronic inflammation, metabolic dysregulation, and immune suppression [12–16]. Visceral fat contributes to metabolic dysfunction by promoting insulin resistance [12] and secreting pro-inflammatory lipids, cytokines, and adipokines (e.g., leptin) [13], which can activate oncogenic pathways such as JAK/STAT and PI3K/AKT [14]. Sarcopenia, characterized by loss of skeletal muscle mass and function, is associated with systemic inflammation and impaired immune surveillance [15], further facilitating tumorigenesis [16].

Despite growing evidence linking body composition to melanoma biology, no studies have leveraged LDCT-based OS to assess melanoma risk, an unmet clinical need, given the high mortality of advanced melanoma.

This study investigates the potential of LDCT-derived body composition features for assessing melanoma risk, using data from the ongoing Pittsburgh Lung Screening Study (PLuSS), initiated in 2002 [17]. We hypothesize that body composition reflects underlying metabolic health and may serve as a valuable resource for melanoma risk assessment. Using our previously developed AI algorithms [18], we segmented and quantified a comprehensive set of three-dimensional (3-D) body composition metrics from baseline LDCT scans in the PLuSS cohort. These measurements were analyzed for their association with melanoma incidence risk using competing risk time-to-event analysis, providing insights into potential biomarkers for melanoma early detection and risk stratification.

## 2. Methods and Materials

2.1. **Study population**

This study used data from the PLuSS cohort [17], which enrolled 3,635 subjects. Inclusion criteria were: (1) age 50 to 79 years; (2) current or former cigarette smokers with a history of smoking at least half a pack per day for 25 years or more; and (3) body weight under 400 pounds. Exclusion criteria were: (1) history of lung cancer; (2) chest CT scan within 12 months before enrollment; and (3) having quit smoking more than 10 years prior to enrollment. Subject data were de-identified and re-identified with unique study IDs by an honest broker. Malignant melanoma diagnoses were confirmed through clinical reports.

As all melanoma cases in this cohort occurred in white participants, the current analysis was restricted to the 3,422 (94.1%) white individuals. At baseline, this subgroup had a mean age of 59.1 years (± 6.2), with 1,776 (51.9%) males (Table 1). Among them, 2,038 (59.6%) were current smokers, and the average BMI was 28.7 (± 5.4). Follow-up time ranged from 0.3 to 21.9 years, with a median of 19.5 years and a mean of 16.8 years. This study was approved by the University of Pittsburgh Institutional Review Board (IRB # 21020128).

**Table 1.** Demographic characteristics, CT-derived body composition summary statistics, and hazard ratios for melanoma incidence (n=3,422).

| Variable | Mean (SD)/Count (%) | Hazard Ratio (95% CI) | P-value |
| --- | --- | --- | --- |
| **Demographics** |  |  |  |
| Age (year) | 59.11 (6.19) | 0.92 (0.75, 1.13) | 0.441 |
| Sex |  |  |  |
| Male | 1,776 (51.90) | 1.08 (0.70, 1.68) | 0.729 |
| Female | 1,646 (48.10) |  |  |
| BMI | 28.71 (5.41) | 1.16 (0.96, 1.41) | 0.121 |
| Smoking Status |  |  |  |
| Current | 2,038 (59.56) | 0.68 (0.44, 1.05) | 0.081 |
| Former | 1,384 (40.44) |  |  |
| **Body Composition** |  |  |  |
| VAT volume (L) | 0.89 (0.59) | 1.24 (1.03, 1.49) | **0.025\*** |
| VAT ratio | 0.08 (0.04) | 1.16 (0.95, 1.43) | 0.150 |
| VAT density (HU) | -90.50 (7.98) | 0.76 (0.63, 0.91) | **0.003\*** |
| SAT volume (L) | 4.13 (1.89) | 1.26 (1.04, 1.52) | **0.016\*** |
| SAT ratio | 0.37 (0.11) | 1.14 (0.92, 1.41) | 0.236 |
| SAT density (HU) | -89.86 (11.55) | 0.80 (0.59, 1.07) | 0.125 |
| IMAT volume (L) | 0.48 (0.27) | 1.18 (1.00, 1.39) | **0.049\*** |
| IMAT ratio | 0.04 (0.02) | 1.10 (0.91, 1.32) | 0.334 |
| IMAT density | -100.59 (13.30) | 0.81 (0.67, 0.99) | **0.036\*** |
| SM volume (L) | 3.98 (1.08) | 1.13 (0.91, 1.39) | 0.274 |
| SM ratio | 0.38 (0.09) | 0.85 (0.68, 1.07) | 0.157 |
| SM density (HU) | 35.01 (7.25) | 0.82 (0.68, 0.99) | **0.039\*** |
| Bone volume (L) | 1.38 (0.32) | 1.04 (0.83, 1.29) | 0.750 |
| Bone ratio | 0.13 (0.03) | 0.78 (0.61, 0.99) | **0.045\*** |
| Bone density (HU) | 324.26 (42.31) | 1.21 (0.94, 1.54) | 0.133 |

Mean (Standard deviation, SD) and Count (%) for continuous and categorical variables respectively.
Hazard ratios and p-values are from univariate time-to-event models. \* indicates p-value < 0.05.
BMI: body mass index, VAT: visceral adipose tissue, SAT: subcutaneous adipose tissue, IMAT: intramuscular SM: skeletal muscle.

### 2.2. Image acquisition

Chest LDCT scans in the PLuSS cohort were acquired over more than 10 years using different protocols and scanner models, including Optima-CT660, LightSpeed-VCT, LightSpeed-Ultra, Siemens Emotion, and Siemens Emotion-Duo. All scans were acquired at end-inspiration during a single breath hold using a helical technique. Images were reconstructed to encompass the entire lung field in a 512×512-pixel matrix with different reconstruction kernels. In-plane pixel dimensions ranged from 0.55 to 0.83 mm, and the slice thickness ranged from 1.25 to 2.5 mm [17].

### 2.3. CT image features

For each patient, a single baseline chest LDCT scan was analyzed. Previously validated AI algorithms [18, 19] were used to automatically segment five body tissue types: visceral adipose tissue (VAT), subcutaneous adipose tissue (SAT), intramuscular adipose tissue (IMAT), skeletal muscle (SM), and bone (Figure S1). These algorithms have been validated in multiple independent studies [20–22]. For each tissue, three quantitative metrics were derived: (1) volume, (2) density, measured as the average Hounsfield Unit (HU) value, and (3) ratio, defined as the tissue volume divided by the total volume of all five segmented tissues within the chest scan.

### 2.4. Demographic and clinical features

Demographic and clinical variables included age (at the time of baseline LDCT), sex, body mass index (BMI), smoking status, and follow-up time from baseline to melanoma diagnosis. Smoking status was categorized as current or former smoker.

### 2.5. Statistical analyses

Fine-Gray subdistribution hazard models were used to analyze the impact of body composition and demographic features on melanoma development, considering death as a competing risk. Both univariate and multivariable analyses were conducted using baseline LDCT scans, with separate analyses for the overall cohort and sex-specific subgroups. Numeric variables were z-score standardized, and categorical variables were encoded as factors. There were no missing data for variables included in the analysis.

To build robust multivariable Fine-Gray models and minimize potential overfitting, feature selection was performed using Least Absolute Shrinkage and Selection Operator (LASSO) regularization, combined with a variance inflation factor (VIF) threshold of < 3. Hazard ratios (HRs) with 95% confidence intervals (CIs) were reported to evaluate the effects of features on melanoma incidence risk. Model performance was evaluated using the concordance index (C-index), receiver operating characteristic (ROC) curves at 5, 10, and 15 years, and the time-dependent area under the curve (AUC) estimates with 95% CIs. Yearly time-dependent AUCs and their 95% CIs were obtained using 5-fold stratified cross-validation. The 21-year average AUC and its 95% CI were computed based on the standard error of the yearly AUCs and the corresponding t-distribution critical value. Additionally, permutation-based importance analysis was conducted to illustrate the relative contribution of each feature to the model’s performance (C-index) for models that retained multiple features.

In addition to hazard-based analyses, restricted mean survival time (RMST) was calculated to evaluate the added prognostic value of body composition metrics beyond traditional BMI. All participants were stratified into high and low groups based on the median values of BMI and each body composition metric, respectively. RMST at 5, 10, and 15 years was then computed to compare average melanoma-free survival time between these groups.

Participants (overall and by sex) were stratified into low-, intermediate-, and high-risk groups based on the 20th and 80th percentiles of the risk predictions from the multivariate models. Cumulative incidence functions were estimated to visually stratify participants by parameters and risk groups. Gray’s test was used to assess the separation of cumulative incidence curves. Statistical significance was defined as p-values < 0.05. Analyses were performed using Python 3.11.4 and RStudio 4.3.1.

## 3. Results

### 3.1. Participant characteristics

As of February 2024, 80 participants in the PLuSS cohort (n=3,635) had been diagnosed with malignant melanoma, all of whom were white. Among white participants (n=3,422), 1,380 deaths occurred, including 4 deaths attributed to melanoma. Time to melanoma diagnosis ranged from 1.0 to 19.9 years, with a median of 9.1 years (interquartile range [IQR]: 5.7-13.7) and a mean of 9.5 years. The cumulative incidence of melanoma was 22.5% (95% CI: 14.0%–32.2%) at 5 years, 53.8% (95% CI: 42.1%–64.0%) at 10 years, and 80.0% (95% CI: 69.2%–87.3%) at 15 years (Figure 1). All subsequent analyses were limited to white participants.

**Figure 1.**
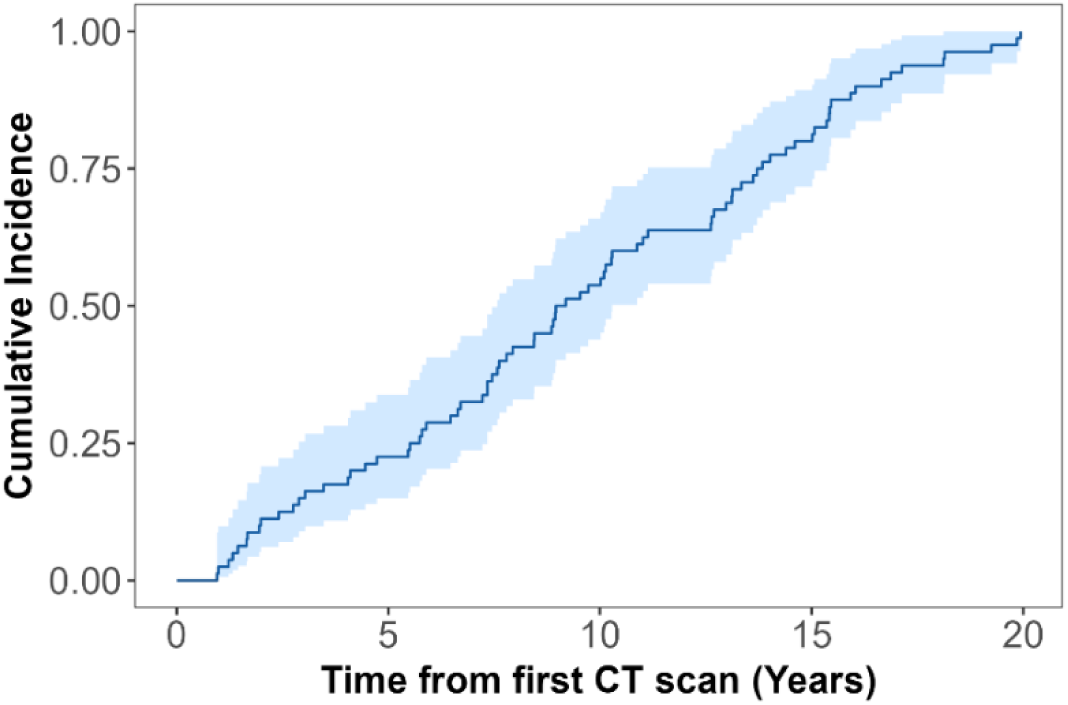
Cumulative incidence function for the melanoma patients (n=80).

Among demographic and clinical variables, none showed statistical significance in univariate analysis, except for *smoking status*, which showed marginal significance (p=0.081, Table 1). Current smokers exhibited a reduced risk of melanoma compared to former smokers (HR=0.68). Cumulative incidence curves stratified by *age group* (< 60 years vs. ≥ 60 years), *sex*, and *smoking status* are presented in Figure 2. Participants aged ≥ 60 years displayed a higher cumulative incidence of melanoma than those aged < 60 (p=0.806) during the first 17 years of follow-up, but this trend reversed beyond 17 years, with younger participants showing higher incidence. Current smokers (p=0.080) and females (p=0.729) consistently exhibited lower cumulative incidence compared to former smokers and males, respectively, throughout the 20-year follow-up period.

**Figure 2.**
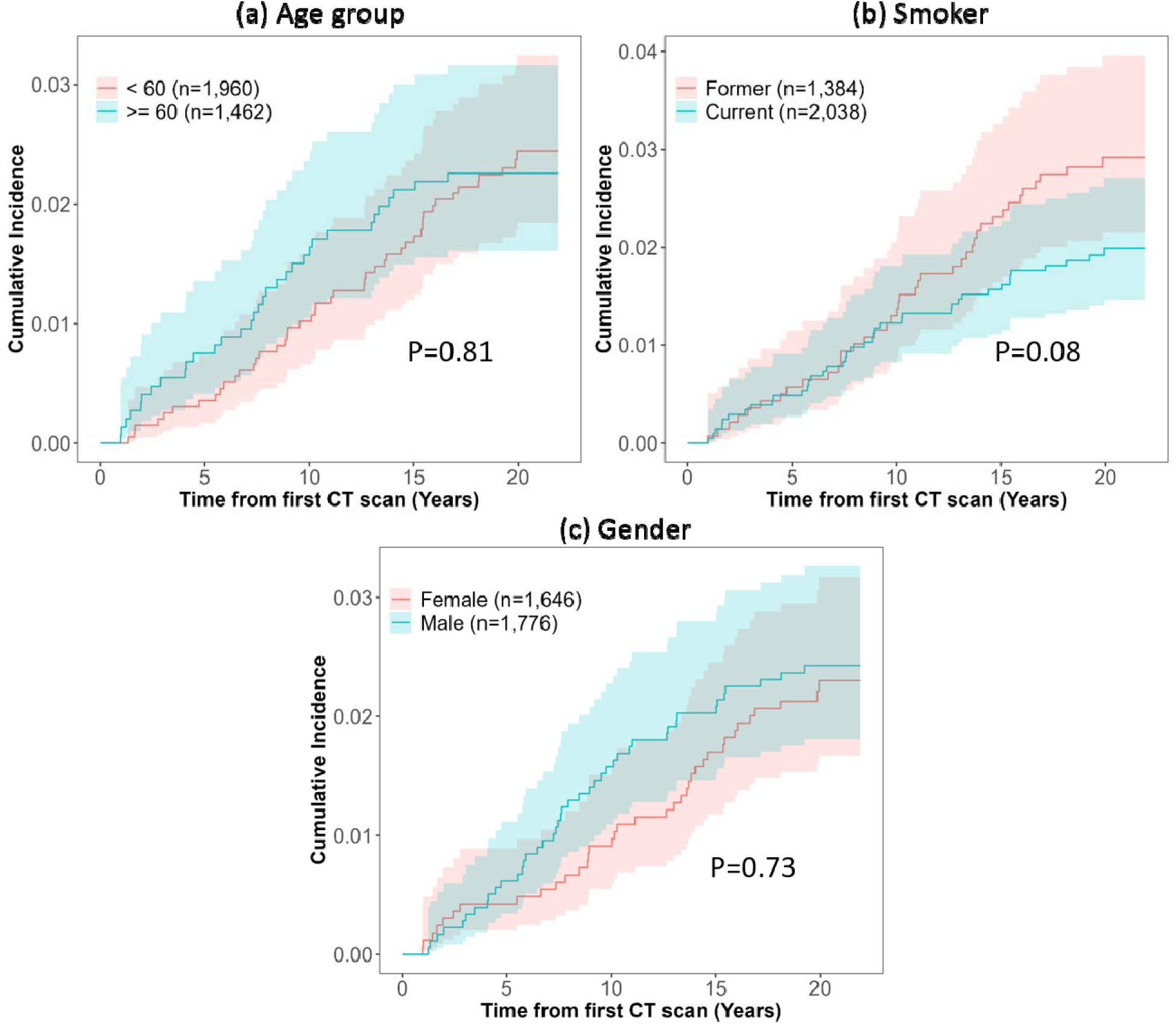
Cumulative incidence function of melanoma patients (n=80) in the PLuSS cohort (n=3,422), stratified by (a) age group, (b) smoking status, and (c) gender.

Among the participants, 43 males and 37 females developed melanoma. Cumulative incidence curves stratified by *age group* (< 60 years vs. ≥ 60 years) and *smoking status* exhibited similar trends when analyzed separately for males and females (Figure S2). Demographic and body composition characteristics by sex are summarized in Tables S1 and S2. Among females, *smoking status* was the only univariately significant predictor of melanoma incidence; current female smokers had a decreased risk compared to former smokers (HR=0.46, p=0.018; Table S2). However, a model relying solely on smoking status demonstrated very limited discriminatory ability in females, with an average 21-year time-dependent AUC of 0.55 (95% CI: 0.53, 0.57; Figure S4).

### 3.2. Association of CT-derived body composition with melanoma incidence

Several body composition features were significantly associated with melanoma incidence in univariate analysis (Table 1). Higher *VAT volume* (HR=1.24, p =0.025), *SAT volume* (HR=1.26, p=0.016), and *IMAT volume* (HR=1.18, p =0.049) were linked to increased risk of melanoma, while higher *VAT density* (HR=0.76, p=0.003), *IMAT density* (HR=0.81, p=0.036), *SM density* (HR=0.82, p=0.039) and *Bone ratio* (HR=0.78, p=0.045) were associated with reduced risk. Sex-specific analyses identified 9 significant body composition features in males (Table S1) and 3 in females (Table S2). In males, increased *SAT volume* (HR=1.29, p=0.020), *SAT ratio* (HR=1.36, p=0.034), *IMAT volume* (HR=1.29, p=0.008), and *IMAT ratio* (HR=1.29, p=0.024) were linked to higher melanoma risk. Conversely, higher *VAT density* (HR=0.75, p=0.004), *IMAT density* (HR=0.74, p=0.027), *SM ratio* (HR=0.72, p=0.040), *SM density* (HR=0.74, p<0.001) and *bone ratio* (HR=0.70, p=0.027) were associated with decreased risk. In females, increased *VAT volume* (HR=1.30, p=0.040), *SM volume* (HR=1.31, p=0.038), and *bone density* (HR=1.58, p=0.010) were associated with increased melanoma risk.

The multivariable models for all participants, males, and females are summarized in Table 2, S3, and S4, respectively. Incorporating demographic features did not improve model performance in any group. For the overall cohort, the final model retained three body composition features—*VAT volume* (HR=1.27, p=0.005), *SM density* (HR=0.81, p=0.044), *and bone density* (HR=1.33, p=0.015)—achieving an average 21-year time-dependent AUC of 0.68 (95% CI: 0.65, 0.70). The model’s AUC was 0.70 at 5 years, 0.67 at 10 years, and 0.65 at 15 years (Figure 3).

**Figure 3.**
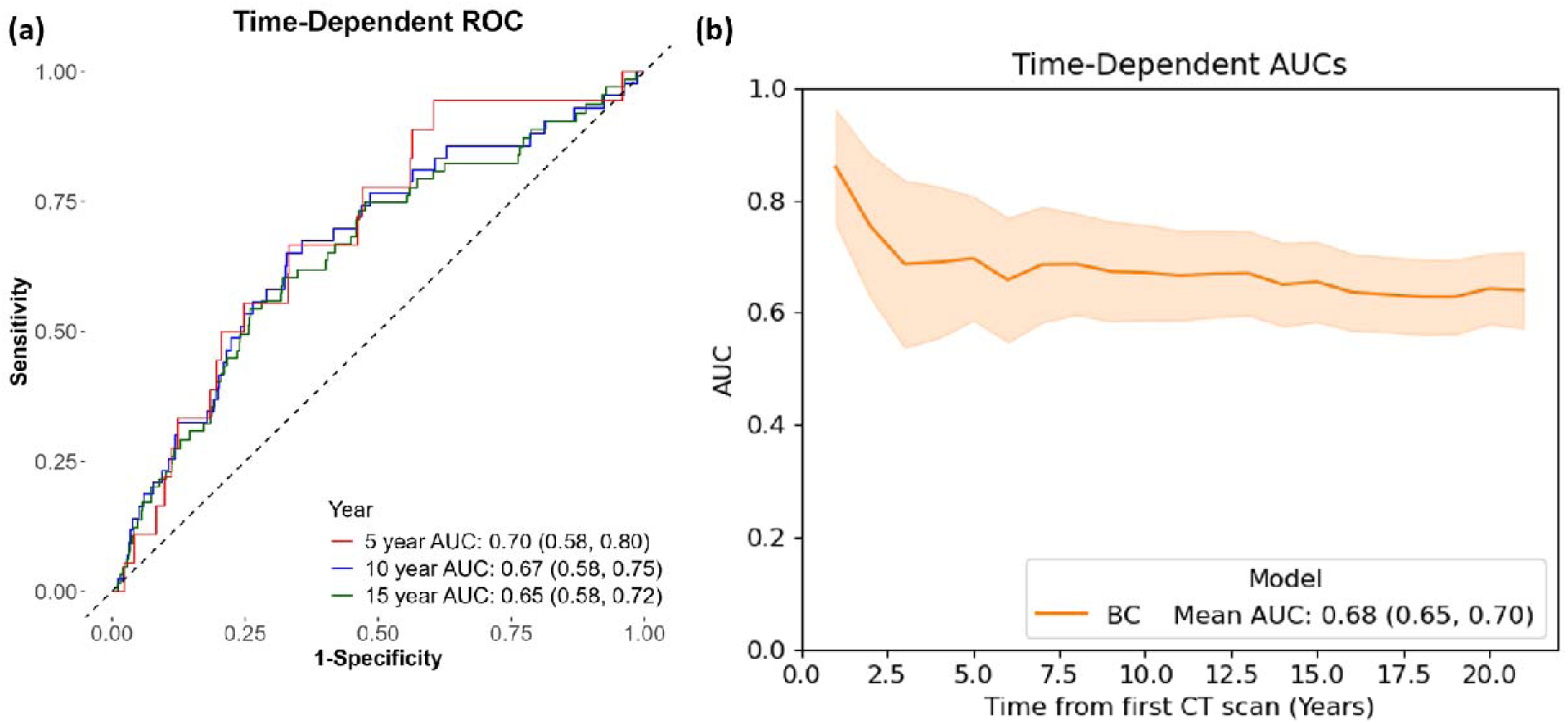
(a) Time-dependent ROC curves of the body composition (BC) model for predicting melanoma incidence at 5, 10, and 15 years. (b) Five-fold cross-validated time-dependent AUCs of the BC model over 21 years.

**Table 2.** Multivariable composite models for predicting melanoma incidence (n=3,422).

| Group | Variables | <u>BC Model</u><br>Hazard Ratio<br>(95% CI) | P-Value |
| --- | --- | --- | --- |
| **Body composition** | VAT volume | 1.27 (1.07, 1.50) | 0.005 |
|  | SM density | 0.81 (0.67, 0.99) | 0.044 |
|  | Bone density | 1.33 (1.06, 1.68) | 0.015 |
| **Concordance index** |  | 0.62 (se = 0.03) |  |
| **AIC** |  | 1291.75 |  |
| Univariate model including *Current smoker*: C-index=0.55 (se=0.03); AIC=1297.55. |  |  |  |
| *BC* — *Body Composition* model. |  |  |  |
| Incorporating demographic features did not improve the predictive performance of the *BC* model. |  |  |  |
| VAT: visceral adipose tissue, SM: skeletal muscle. |  |  |  |

For males, *SM density* (HR=0.74, p<0.001) was the only retained feature, yielding an average 21-year time-dependent AUC of 0.67 (95% CI: 0.65, 0.68), and AUCs of 0.71, 0.63, and 0.67 at 5, 10, and 15 years, respectively (Figure S3). For females, the model included *VAT volume* (HR=1.47, p<0.0001), *IMAT ratio* (HR=0.67, p=0.019), and *bone density* (HR=1.75, p=0.002), achieving an average 21-year time-dependent AUC of 0.68 (95% CI: 0.65, 0.71), with AUCs of 0.62, 0.72, and 0.67 at 5, 10, and 15 years, respectively (Figure S4). Both sex-specific models satisfied the proportional subdistribution hazards assumption (Table S5). Permutation-based feature importance, measured by the C-index, is illustrated in Figure S5. In both the overall and female models, *VAT volume* ranked as the most important feature.

### 3.3. Restricted mean survival time (RMST) comparison

Participants were stratified into high and low groups based on the median values of BMI, adipose tissue volumes (*VAT, SAT, IMAT*), *SM density*, and *bone density*. RMST analyses at 5, 10, and 15 years showed that certain body composition metrics provided stronger prognostic stratification than BMI (Table 3). Specifically, higher *VAT volume*, higher *IMAT volume*, and lower *SM density* were consistently associated with numerically shorter melanoma-free survival across all time points and demonstrated better stratification than BMI (Figure 4). *SAT volume* exhibited a similar stratification trend as BMI but did not reach statistical significance. In contrast, *bone density* showed negligible RMST differences between high and low groups, indicating limited discriminatory capacity in this context.

**Figure 4.**
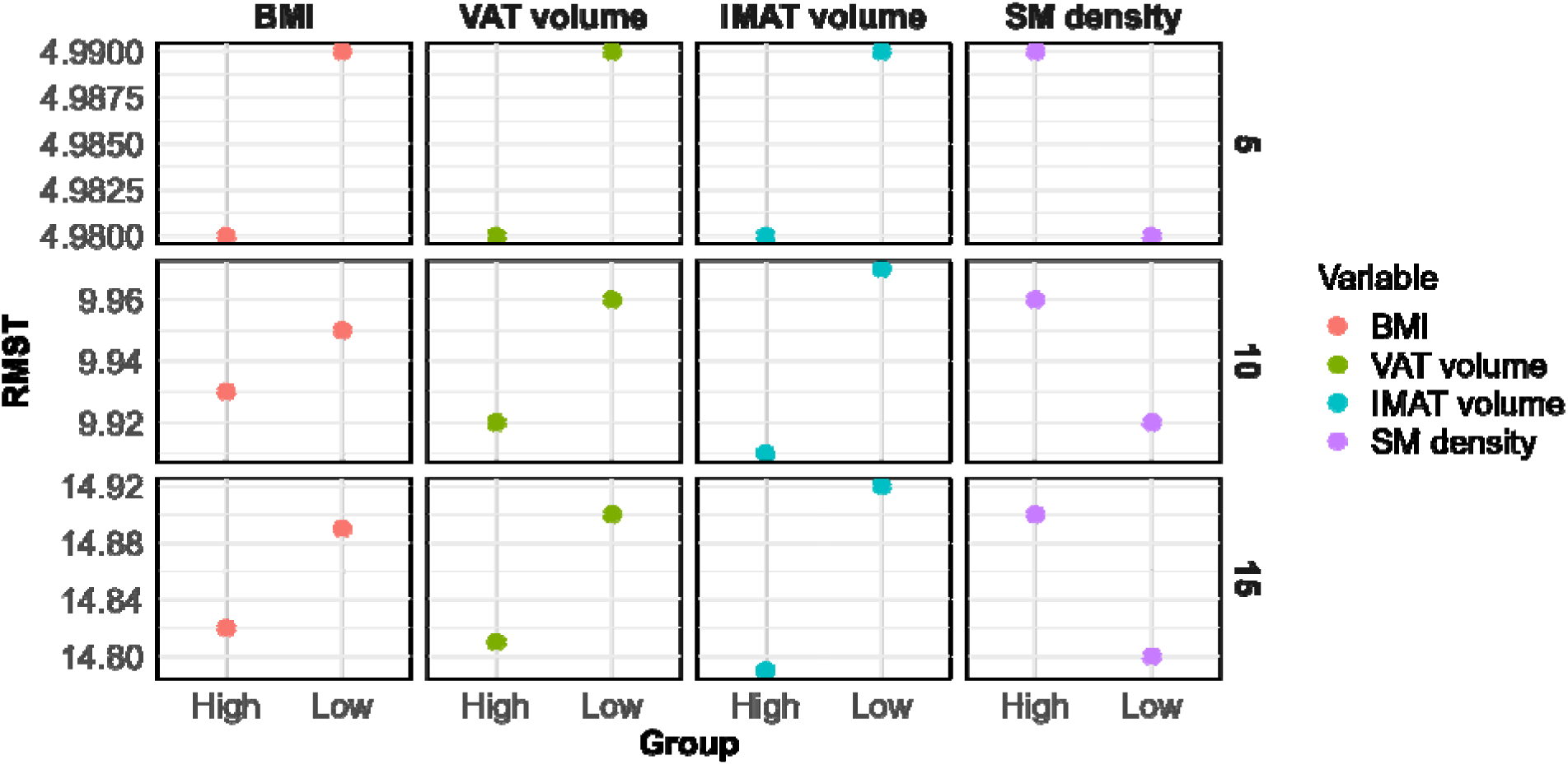
Restricted mean survival time (RMST) comparisons at 5, 10, and 15 years across BMI and body composition metrics.

**Table 3.** Restricted mean survival time (RMST) at 5, 10, and 15 years stratified by body composition and BMI groups for all participants (n=3,422).

| Variable | Group | RMST (5y),<br>(95% CI) | RMST (10y),<br>(95% CI) | RMST (15y),<br>(95% CI) |
| --- | --- | --- | --- | --- |
| BMI | High (>27.92) | 4.98 (4.97, 5.00) | 9.93 (9.90, 9.96) | 14.82 (14.76, 14.88) |
|  | Low (<= 27.92) | 4.99 (4.98, 5.00) | 9.95 (9.92, 9.98) | 14.89 (14.83, 14.94) |
|  | P Value (difference) | 0.504 | 0.358 | 0.124 |
| VAT volume | High (> 0.77 L ) | 4.98 (4.97, 4.99) | 9.92 (9.88, 9.95) | 14.81 (14.74, 14.87) |
|  | Low (<= 0.77 L) | 4.99 (4.98, 5.00) | 9.96 (9.94, 9.98) | 14.90 (14.86, 14.95) |
|  | P Value (difference) | 0.119 | **0.039** | **0.020** |
| SAT volume | High (> 3.80 L ) | 4.98 (4.97, 4.99) | 9.93 (9.89, 9.96) | 14.82 (14.76, 14.88) |
|  | Low (<= 3.80 L) | 4.99 (4.98, 5.00) | 9.95 (9.93, 9.98) | 14.89 (14.84, 14.94) |
|  | P Value (difference) | 0.334 | 0.173 | 0.078 |
| IMAT volume | High (> 0.42 L ) | 4.98 (4.97, 4.99) | 9.91 (9.87, 9.95) | 14.79 (14.72, 14.86) |
|  | Low (<= 0.42 L) | 4.99 (4.99, 5.00) | 9.97 (9.95, 9.99) | 14.92 (14.88, 14.96) |
|  | P Value (difference) | **0.024** | **0.004** | **0.002** |
| SM density | High (> 35.41 HU) | 4.99 (4.99, 5.00) | 9.96 (9.94, 9.98) | 14.90 (14.86, 14.95) |
|  | Low (<= 35.41 HU) | 4.98 (4.97, 4.99) | 9.92 (9.88, 9.95) | 14.80 (14.74, 14.87) |
|  | P Value (difference) | **0.039** | **0.026** | **0.019** |
| Bone density | High (> 322.56 HU) | 4.99 (4.98, 5.00) | 9.94 (9.91, 9.97) | 14.85 (14.79, 14.91) |
|  | Low (<= 322.56 HU) | 4.99 (4.98, 5.00) | 9.94 (9.91, 9.97) | 14.86 (14.80, 14.91) |
|  | P Value (difference) | 0.954 | 0.939 | 0.869 |
| P-values indicate the difference between high and low groups. |  |  |  |  |

**Table 4.** Risk stratification based on the composite model for all participants (n=3,422).

| Risk strata | Melanoma Incidence | Total (%) | Risk estimates (95% CI) |  |  |
| --- | --- | --- | --- | --- | --- |
|  |  |  | 5-year | 10-year | 15-year |
| Low | 8 | 685 (20.0) | 0.15%<br>(0.02, 0.81) | 0.80%<br>(0.31, 1.80) | 1.20%<br>(0.53, 2.40) |
| Intermediate | 46 | 2,052 (60.0) | 0.45%<br>(0.22, 0.83) | 1.10%<br>(0.74, 1.70) | 1.90%<br>(1.40, 2.60) |
| High | 26 | 685 (20.0) | 1.20%<br>(0.57, 2.30) | 2.50%<br>(1.50, 3.90) | 3.60%<br>(2.30, 5.30) |
| Total | 80 | 3,422 |  |  |  |

### 3.4. Risk stratification

Based on the 20th and 80th percentiles of risk prediction scores, participants were stratified into low-, intermediate-, and high-risk groups for the overall cohort, males, and females according to their individual final model predictions. The cumulative incidence functions for these stratifications are visualized in Figure 5 and Figure S6. Tables 4, S6, and S7 summarize the number of melanoma incidence, total participants, and the estimated cumulative incidence within each risk stratum for all participants, males, and females, respectively. Across all time points, males consistently exhibited higher estimated melanoma risks than females in corresponding risk groups. Specifically, in the high-risk group, the estimated risks for males were 1.50%, 3.50%, and 4.70% at 5, 10, and 15 years, respectively. In contrast, females in the high-risk group had estimated risks of 0.91%, 2.20%, and 3.60% at the same time points.

**Figure 5.**
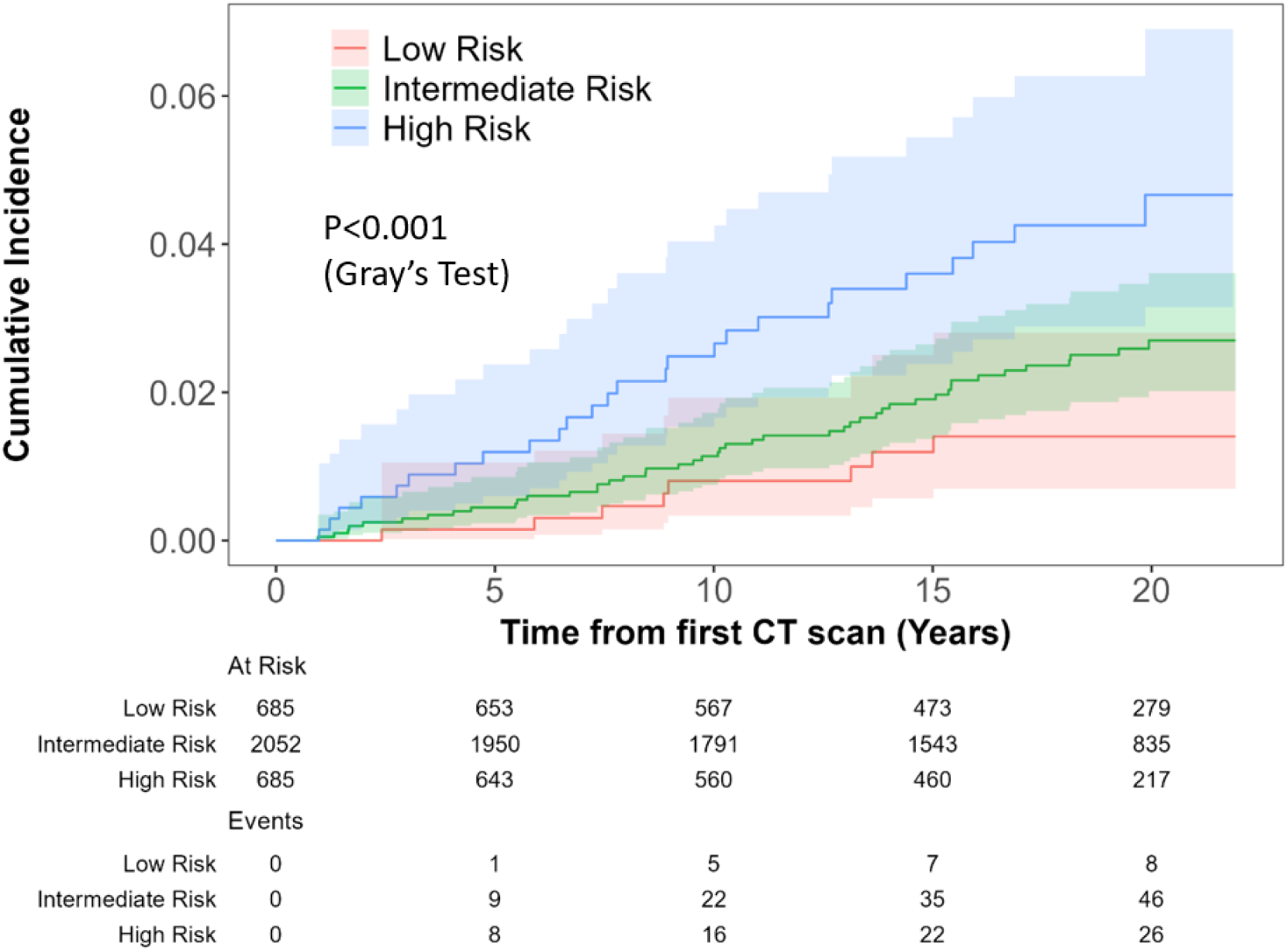
Cumulative incidence of melanoma stratified by low, intermediate, and high-risk groups. Risk stratification is based on the multivariate models for all participants (n=3,422).

## 4. Discussion

LDCT is widely adopted for early lung cancer screening, offering opportunistic screening for evaluating melanoma risk through body composition analysis. In this study, we applied previously developed AI algorithms to segment and quantify five body tissue types from baseline LDCT scans in the PLuSS cohort, evaluating their associations with developing malignant melanoma in the context of lung cancer screening. Given the high competing risks of lung cancer or other cardiovascular-related deaths in the lung cancer screening population, we employed Fine-Gray subdistribution hazard models to account for these events, providing more accurate risk estimates than traditional cause-specific models. Our findings show that CT-derived body composition is significantly associated with melanoma risk and can be potentially used to stratify subjects into different risk groups. While this study focused on screening LDCT scans, this approach could also be extended to diagnostic CTs, enabling the extraction of body composition metrics as a secondary output.

Previous studies have investigated the relationship between BMI and melanoma outcomes [23–28], including incidence, disease-free survival (DFS), progression-free survival (PFS), and overall survival (OS). Findings have been inconsistent: some report a potential increased melanoma risk with higher BMI [24, 26], others find no clear association [27], while some suggest an “obesity paradox”, where higher BMI is associated with better melanoma outcomes [25, 28]. These conflicting results highlight the limitations of BMI as a reliable risk factor. Our study aligns with this pattern, as BMI was not significantly associated with melanoma incidence in the overall cohort or in sex-specific analyses.

Imaging-derived body composition analysis has emerged as a powerful tool to overcome the limitations of BMI by providing a comprehensive evaluation of body composition. However, most studies have focused on the prognostic value of body composition in melanoma patients undergoing treatment [29–31], leaving its association with melanoma incidence risk unexplored. Our study fills this gap, demonstrating that CT-derived body composition features were associated with melanoma incidence risk in the overall cohort (Tables 1 and 2) as well as in sex-specific analyses (Tables S1-S4).

In males, higher adipose tissue volumes and ratios, including *SAT* and *IMAT*, were linked to higher melanoma risk, while greater *SM ratio*, *SM density,* and *bone ratio* were associated with reduced risk. In females, higher *VAT volume*, *SM volume*, and *bone density* were linked to higher melanoma risk. These sex-specific discrepancy aligns with previous studies, where overweight and obesity, as measured by BMI, and in our study, by image-derived adiposity, were associated with increased melanoma risk in males [24], while findings in females were inconsistent [23]. Additionally, in females, *SM volume* alone may not fully capture muscle quality or distribution, and its predictive value might be limited by the smaller number of melanoma events.

Additionally, RMST analyses confirmed the added prognostic value of body composition metrics beyond BMI (Table 3, Figure 4). Higher adipose tissue volumes and lower *SM density* were consistently associated with shorter melanoma-free survival, reinforcing the hazard-based findings (Table 1). While *bone density* was included in multivariate models for females and the overall cohort, RMST results suggest it may not be a strong independent predictor, possibly due to sex-specific discrepancies (Tables S1, S2).

The multivariate model demonstrated promising performance, with the all-participant model achieving an average 21-year AUC of 0.68 (Figure 3). Model performance was highest during the first 2.5 years, followed by a decline and subsequent stabilization. The male-specific model, which included only *SM density*, achieved an average 21-year AUC of 0.67 (Figure S3), with improved performance in the first 5 years before decreasing and stabilizing. Similarly, the female-specific model exhibited a similar trend to the all-participant model, peaking in the first 2.5 years, with an average 21-year AUC of 0.68 (Figure S4). When comparing AUCs at 5, 10, and 15 years, the all-participant and male models performed the best at 5 years, while the female model achieved its highest performance at 10 years. These results collectively highlight the predictive capabilities of our models in both short-term and long-term contexts.

Many studies have shown that melanoma risk increases with age [6, 32–34], except among young adults, where melanoma remains one of the most common cancers in individuals under 30 [6, 34]. Additionally, male sex has been associated with a higher melanoma risk, especially after age 50 [35, 36]. However, neither age nor sex was found to be significantly predictive in the PLuSS cohort (Table 1, S1, S2). Potential reasons for this could be: (1) the relatively small number of melanoma incidence in the PLuSS cohort (80 out of 3,422), and (2) the narrower age range of 50 to 70 years in the PLuSS, compared to other studies that encompass a wider age range (e.g., 0 to 90 years) [33, 34]. Although age and sex were not statistically significant, cumulative incidence functions (Figure 2) revealed trends suggesting higher melanoma risk in older individuals and males. Interestingly, when stratified by age group, the cumulative incidence in the older group decreased after long-term follow-up (>17 years). This decline is likely due to the higher impact of competing risks, such as deaths from other causes, in the older group.

Smoking status also showed noteworthy patterns. Although its predictive power was limited (time-dependent AUC= 0.55 among females; Figure S4), current smoking was inversely associated with melanoma incidence compared to former smoking (Table 1, S1, and S2; Figure 2 and S2). This finding aligns with previous studies reporting a reduced melanoma risk among current or former smokers compared to non-smokers [37–39], suggesting a consistent inverse association between cigarette smoking and melanoma risk.

Our validated AI algorithm can automatically extract body composition metrics from routine CT scans, enabling opportunistic melanoma risk assessment without additional imaging or workflow changes. For clinical implementation, further validation of our findings in diverse populations, integration with existing PACS systems, and alignment with personalized care pathways will be essential.

The primary limitation of this study is the lack of an external dataset for validation, which may limit generalizability. However, as the first study to explore the relationship between imaging-derived body composition and melanoma risk in the context of lung cancer screening, it offers novel insights. Given the study’s focus on opportunistic use of lung screening data, some limitations are inherent to the design. Key melanoma risk factors, such as UV exposure and number of nevi, were unavailable in the PLuSS cohort, as data collection was not tailored for melanoma risk assessment. Additionally, the cohort included only older current or former smokers, limiting applicability to younger or non-smoker populations. However, melanoma incidence is relatively low among individuals under 50 years old (∼7.8% worldwide annually [6]), and most cases occur in older adults, aligning our study with the highest-risk group. Future studies should aim to validate these findings in more diverse populations, including never smokers.

## 5. Conclusion

We evaluated low-dose CT scans from lung cancer screening as a potential tool for melanoma incidence assessment by analyzing CT-derived body composition and demographic characteristics. Competing risk time-to-event analysis was performed. Our investigation showed that melanoma risk was associated with CT-derived body composition, with males exhibiting higher estimated risk compared to females in both the short-term and long-term. Given melanoma’s high mortality and the limitations of current screening programs, this study highlights the opportunity to leverage widely available lung cancer screening to ultimately complement existing methods, enhance early detection and improve melanoma risk assessment.

## Data Availability

All data produced in the present study are available upon reasonable request to the authors

## Abbreviations

AI: artificial intelligence
BC: body composition
CI: confidence interval
HR: hazard ratio
HU: Hounsfield unit
IMAT: intramuscular adipose tissue
LDCT: low-dose computed tomography
OS: opportunistic screening
ROC-AUC: the area under the receiver operating characteristic curve
SAT: subcutaneous adipose tissue
SM: skeletal muscle
VAT: visceral adipose tissue

