## Supplemental Material for "Body composition and melanoma incidence risk: insights from a longitudinal lung cancer screening cohort"

**Table S1.** Demographic characteristics, CT-derived body composition summary statistics, and hazard ratios for melanoma incidence/Males (n=1,776).

| Variable | | | Mean (SD)/Count (%) | Hazard Ratio (95% CI) | P-value |
| --- | --- | --- | --- | --- | --- |
| Demographics | | | | | |
| Age (year) | | | 59.30 (6.25) | 1.03 (0.78, 1.37) | 0.823 |
| BMI | | | 29.25 (4.94) | 1.13 (0.88, 1.46) | 0.335 |
| Smoking Status |  | |  |  |  |
|  | Current | | 1,011 (56.93) | 0.96 (0.52, 1.74) | 0.881 |
|  | Former | | 765 (43.07) |  |  |
| Body Composition | | | | | |
| VAT volume (L) | |  | 1.19 (0.61) | 1.22 (0.95, 1.57) | 0.113 |
| VAT ratio | |  | 0.09 (0.04) | 1.15 (0.88, 1.50) | 0.300 |
| VAT density (HU) | |  | -92.01 (7.86) | 0.75 (0.61, 0.91) | **0.004^*^** |
| SAT volume (L) | |  | 3.80 (1.73) | 1.29 (1.04, 1.59) | **0.020^*^** |
| SAT ratio | |  | 0.30 (0.08) | 1.36 (1.02, 1.80) | **0.034** |
| SAT density (HU) | |  | -87.56 (12.16) | 0.67 (0.44, 1.01) | 0.058 |
| IMAT volume (L) | |  | 0.55 (0.29) | 1.29 (1.07, 1.54) | **0.008^*^** |
| IMAT ratio | |  | 0.04 (0.02) | 1.29 (1.03, 1.61) | **0.024^*^** |
| IMAT density (HU) | |  | -103.19 (14.19) | 0.74 (0.56, 0.97) | **0.027^*^** |
| SM volume (L) | |  | 4.83 (0.75) | 1.08 (0.78, 1.50) | 0.649 |
| SM ratio | |  | 0.41 (0.08) | 0.72 (0.53, 0.98) | **0.040^*^** |
| SM density (HU) | |  | 36.78 (6.94) | 0.74 (0.62, 0.88) | **<0.001^**^** |
| Bone volume (L) | |  | 1.63 (0.20) | 0.99 (0.70, 1.40) | 0.956 |
| Bone ratio | |  | 0.14 (0.03) | 0.70 (0.51, 0.96) | **0.027^*^** |
| Bone density (HU) | |  | 314.13 (37.49) | 0.94 (0.69, 1.27) | 0.681 |
| Mean (Standard deviation, SD) and Count (%) for continuous and categorical variables respectively. | | | | | |
| ^*^, ^**^, indicate p-value < 0.05, < 0.001. | | | | | |
| BMI: body mass index, VAT: visceral adipose tissue, SAT: subcutaneous adipose tissue, IMAT: intramuscular adipose tissue, | | | | | |
| SM: skeletal muscle. | | | | | |

**Table S2.** Demographic characteristics, CT-derived body composition summary statistics, and hazard ratios for melanoma incidence/Females (n=1,646).

| Variable | | Mean (SD)/Count (%) | Hazard Ratio (95% CI) | P-value |
| --- | --- | --- | --- | --- |
| Demographics | | | | |
| Age (year) | | 58.90 (6.11) | 0.80 (0.60, 1.07) | 0.132 |
| BMI | | 28.12 (5.82) | 1.19 (0.89, 1.58) | 0.239 |
| Smoking Status |  |  |  |  |
|  | Current | 1,027 (62.39) | 0.46 (0.24, 0.88) | **0.018** |
|  | Former | 619 (37.61) |  |  |
| Body Composition | | | | |
| VAT volume (L) |  | 0.57 (0.36) | 1.30 (1.01, 1.68) | **0.040^*^** |
| VAT ratio |  | 0.06 (0.02) | 1.21 (0.84, 1.74) | 0.304 |
| VAT density (HU) |  | -88.87 (7.79) | 0.82 (0.64, 1.04) | 0.099 |
| SAT volume (L) |  | 4.48 (1.99) | 1.25 (0.92, 1.72) | 0.159 |
| SAT ratio |  | 0.45 (0.09) | 1.11 (0.77, 1.59) | 0.587 |
| SAT density (HU) |  | -92.34 (10.28) | 0.95 (0.71, 1.28) | 0.756 |
| IMAT volume (L) |  | 0.40 (0.22) | 1.00 (0.75, 1.33) | 0.980 |
| IMAT ratio |  | 0.04 (0.01) | 0.81 (0.60, 1.10) | 0.179 |
| IMAT density (HU) |  | -97.79 (11.65) | 0.94 (0.75, 1.19) | 0.619 |
| SM volume (L) |  | 3.07 (0.48) | 1.31 (1.02, 1.68) | **0.038^*^** |
| SM ratio |  | 0.34 (0.08) | 0.93 (0.65, 1.34) | 0.713 |
| SM density (HU) |  | 33.11 (7.09) | 1.01 (0.75, 1.36) | 0.940 |
| Bone volume (L) |  | 1.10 (0.14) | 1.04 (0.78, 1.38) | 0.802 |
| Bone ratio |  | 0.12 (0.03) | 0.82 (0.55, 1.23) | 0.348 |
| Bone density (HU) |  | 335.18 (44.45) | 1.58 (1.12, 2.24) | **0.010^*^** |
| Mean (Standard deviation, SD) and Count (%) for continuous and categorical variables respectively. | | | | |
| ^*^ indicates p-value < 0.05. | | | | |
| BMI: body mass index, VAT: visceral adipose tissue, SAT: subcutaneous adipose tissue, IMAT: intramuscular adipose tissue, | | | | |
| SM: skeletal muscle. | | | | |

**Table S3.** Multivariate model for predicting melanoma incidence/Males (n=1,776).

| **Group** | **Variables** | **BC Model** | |
| --- | --- | --- | --- |
|  |  | **Hazard Ratio**  **(95% CI)** | **P-Value** |
| **Body composition** | SM density | 0.74 (0.62, 0.88) | <0.001 |
| **Concordance index** |  | 0.60 (se = 0.04) |  |
| **AIC** |  | 637.38 |  |
| *BC* — *Body Composition* model. | | | |
| Incorporating demographic features did not improve the predictive performance of the *BC* model. | | | |
| SM: skeletal muscle. | | | |

**Table S4.** Multivariate model for predicting melanoma incidence/Females (n=1,646).

| **Group** | **Variables** | **BC Model** | |
| --- | --- | --- | --- |
|  |  | **Hazard Ratio**  **(95% CI)** | **P-Value** |
| **Body composition** | VAT volume | 1.47 (1.21, 1.78) | <0.001 |
|  | IMAT ratio | 0.67 (0.47, 0.94) | 0.019 |
|  | Bone density | 1.75 (1.23, 2.47) | 0.002 |
| **Concordance index** |  | 0.71 (se = 0.04) |  |
| **AIC** |  | 532.74 |  |
| Univariate model including *Current smoker*: C-index=0.60 (se=0.04); AIC=542.16.  *BC* — *Body Composition* model. | | | |
| Incorporating demographic features did not improve the predictive performance of the *BC* model. | | | |
| VAT: visceral adipose tissue, IMAT: intramuscular adipose tissue. | | | |

**Table S5**. Results of the proportional subdistribution hazards assumption test for variables included in the multivariable Fine-Gray models.

| Variables | $\chi^{2}$ | P Value |
| --- | --- | --- |
| Male model | | |
| SM density | 0.648 | 0.420 |
| Female model | | |
| VAT volume | 2.250 | 0.130 |
| IMAT ratio | 1.790 | 0.180 |
| Bone density | 2.110 | 0.150 |
| VAT: visceral adipose tissue; IMAT: intramuscular adipose tissue;  SM: skeletal muscle. | | |

**Table S6.** Risk stratification based on the composite model for male participants (n=1,776).

| **Risk strata** | **Melanoma**  **Incidence** | **Total (%)** | **Risk estimates (95% CI)** | | |
| --- | --- | --- | --- | --- | --- |
|  |  |  | **5-year** | **10-year** | **15-year** |
| Low | 4 | 356 (20.0) | 0.00%  (-, -) | 0.85%  (0.24, 2.3) | 1.20%  (0.40, 2.90) |
| Intermediate | 25 | 1,065 (60.0) | 0.58%  (0.25, 1.20) | 1.30%  (0.78, 2.20) | 2.00%  (1.20, 3.00) |
| High | 14 | 355 (20.0) | 1.50%  (0.56, 3.30) | 3.50%  (1.8, 5.90) | 4.70%  (2.60, 7.70) |
| Total | 43 | 1,776 |  |  |  |

**Table S7.** Risk stratification based on the composite model for female participants (n=1,646).

| **Risk strata** | **Melanoma**  **Incidence** | **Total (%)** | **Risk estimates (95% CI)** | | |
| --- | --- | --- | --- | --- | --- |
|  |  |  | **5-year** | **10-year** | **15-year** |
| Low | 3 | 330 (20.0) | 0.31%  (0.03, 1.60) | 0.31%  (0.03, 1.60) | 1.10%  (0.30, 3.00) |
| Intermediate | 17 | 987 (60.0) | 0.31%  (0.09, 0.85) | 0.74%  (0.33, 1.50) | 1.60%  (0.90, 2.60) |
| High | 17 | 329 (20.0) | 0.91%  (0.26, 2.50) | 2.20%  (0.98, 4.30) | 3.60%  (1.90, 6.10) |
| Total | 37 | 1,646 |  |  |  |

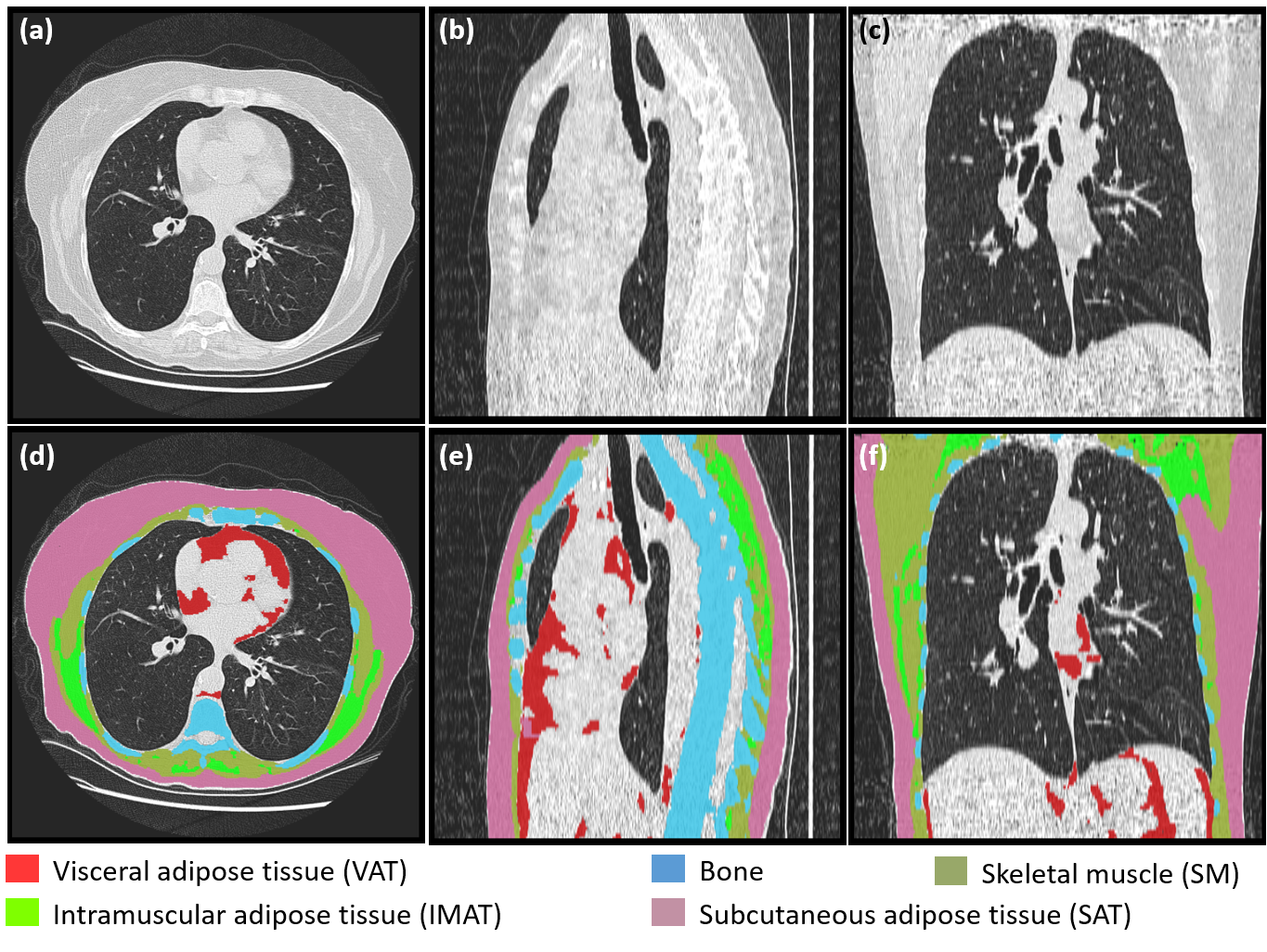

**Figure S1.** Automated segmentations of five tissues depicted on an LDCT scan. (a-c) Axial, coronal, and sagittal images, respectively; (d-f) corresponding segmented images.

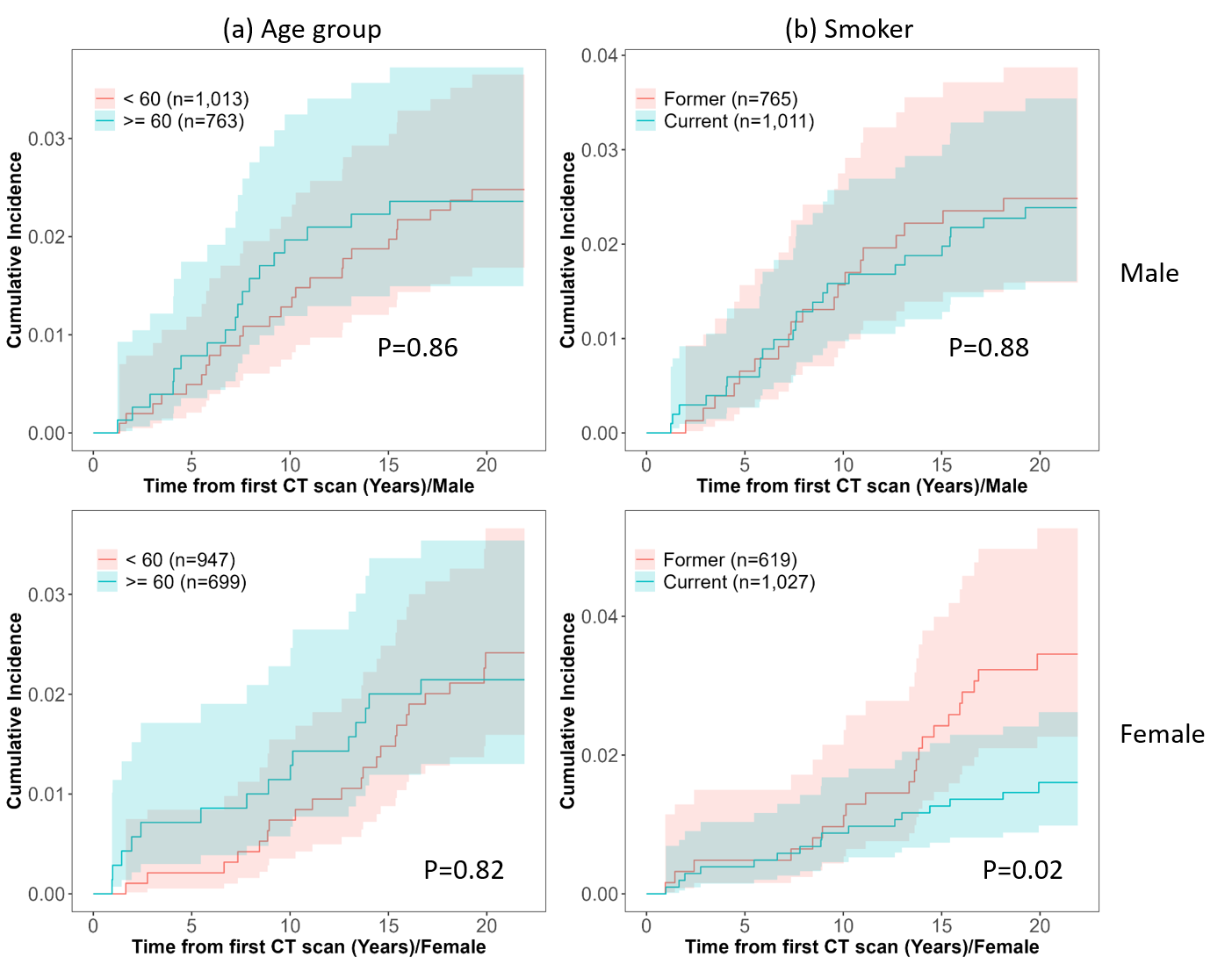

**Figure S2.** Cumulative incidence of melanoma among male patients (n=43) and female patients (n=37) in the PLuSS cohort (n=3,422), stratified by (a) age group and (b) smoking status.

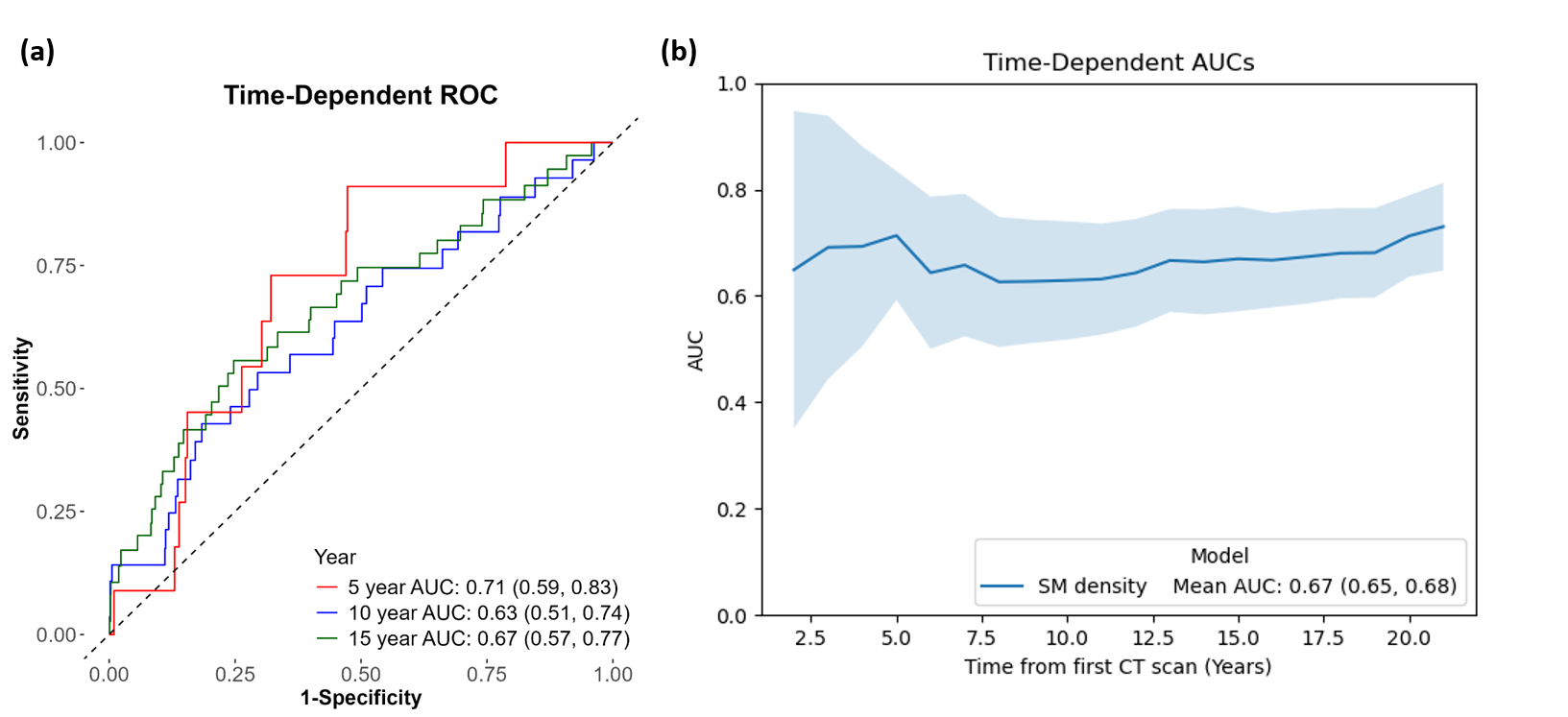

**Figure S3.** (a) Time-dependent ROC curves of the body composition (BC) model (*SM density*) for predicting melanoma incidence at 5, 10, and 15 years among male participants. (b) Five-fold cross-validated time-dependent AUCs of the BC model (*SM density*) over 21 years among male participants (n=1,776).

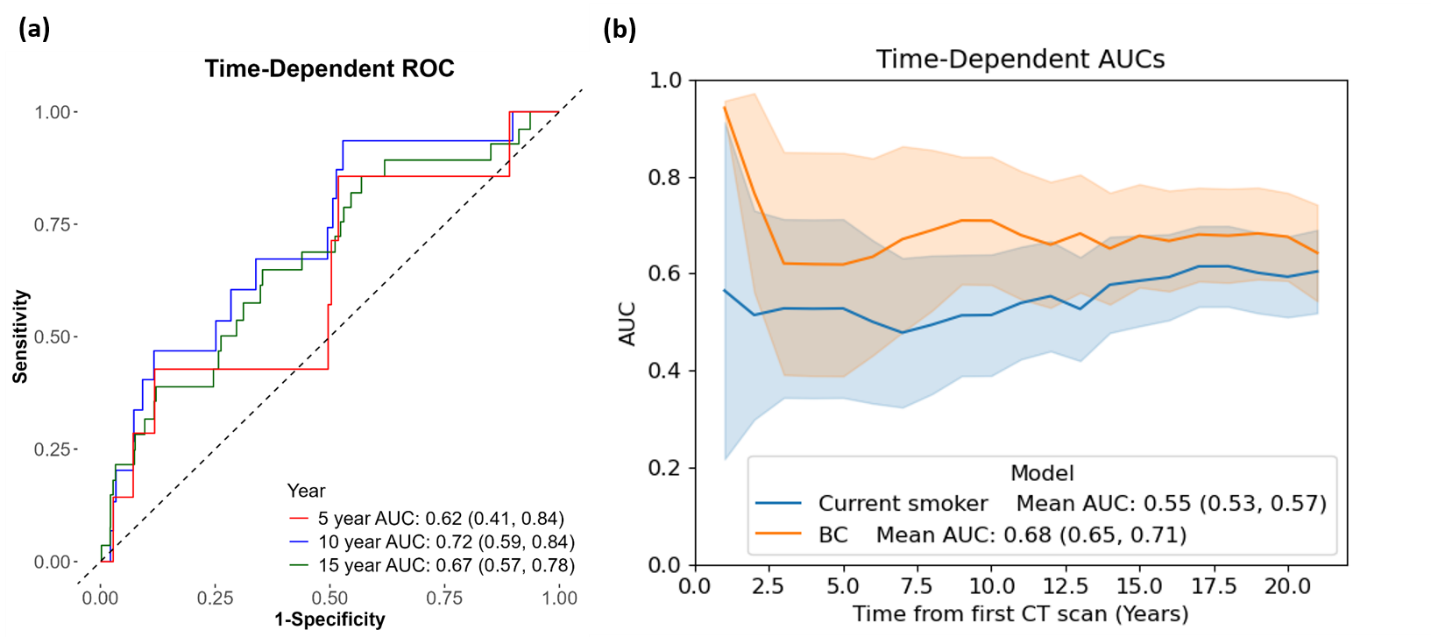

**Figure S4.** (a) Time-dependent ROC curves of the multivariate body composition (BC) model for predicting melanoma incidence at 5, 10, and 15 years among female participants. (b) Five-fold cross-validated time-dependent AUCs of the BC model and the univariate *Current smoker* model over 21 years among female participants (n=1,646).

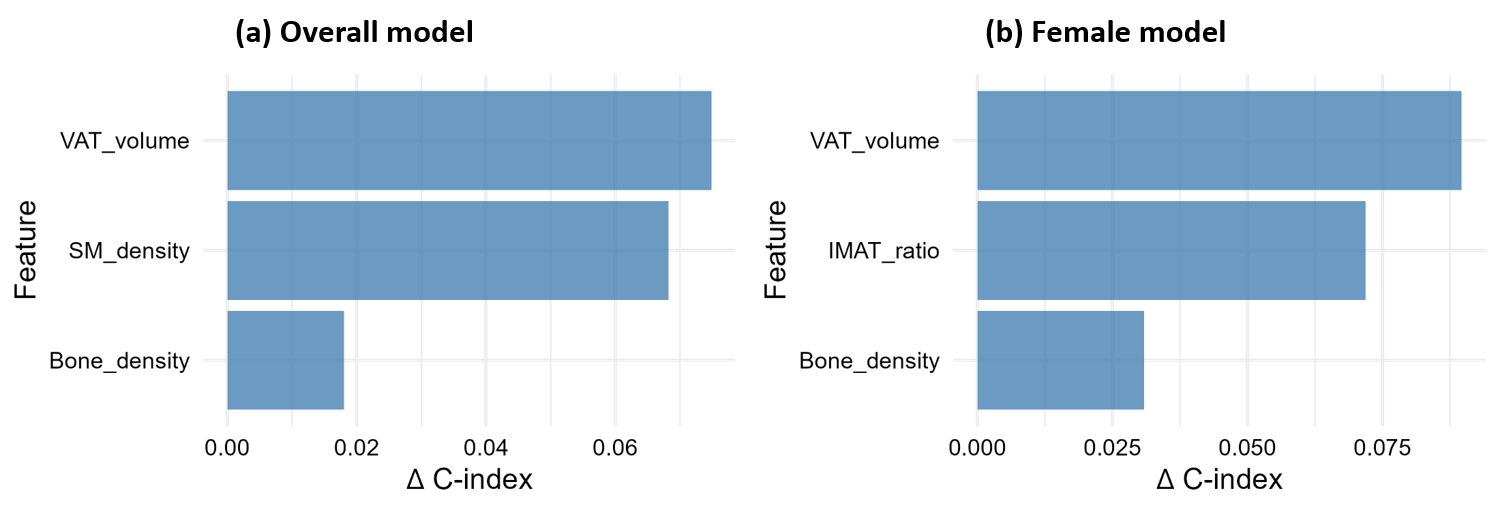

**Figure S5.** Feature importance based on C-Index reduction from permutation: overall and female subgroup models.

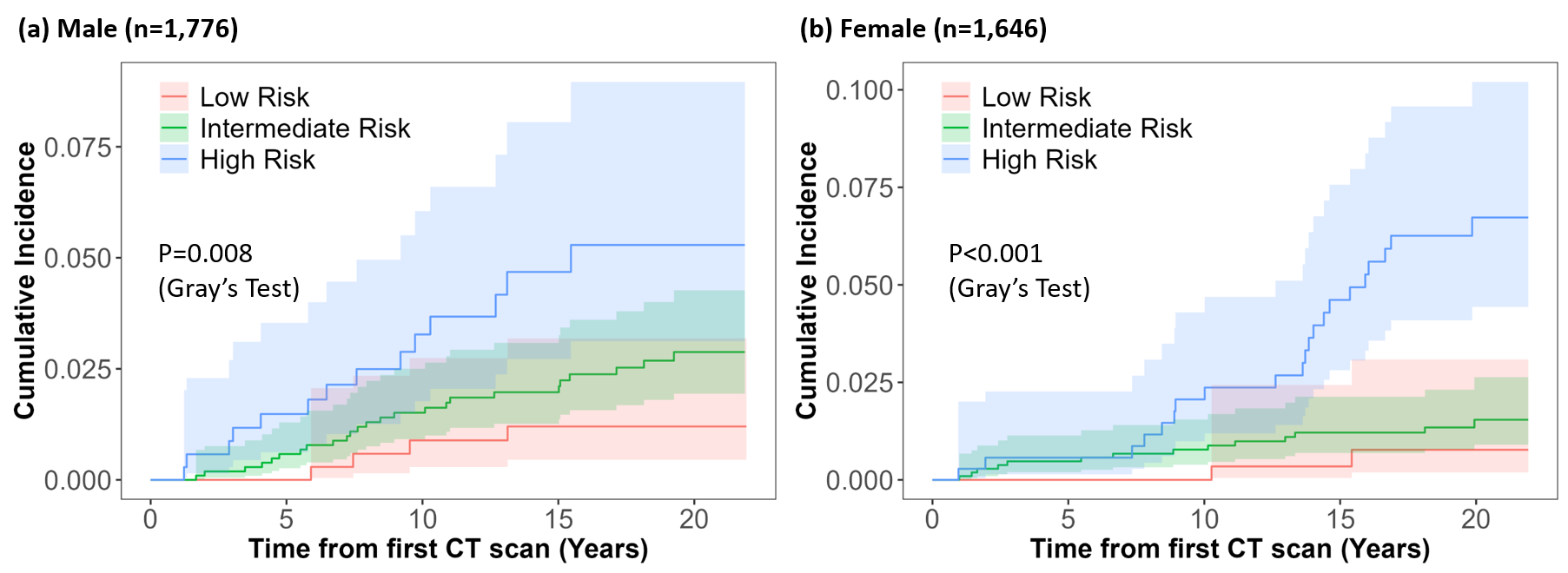

**Figure S6.** Cumulative incidence of melanoma stratified by low, intermediate, and high-risk groups. Risk stratification is based on the composite models for both male (a, n=1,776) and female (b, n=1,646) participants.
